# TCA-soluble blood serum proteins of COVID-19 patients as possible predictive markers for the disease severity

**DOI:** 10.1101/2021.04.07.21255063

**Authors:** Andrii Orfin, Marina Starykovych, Tamila Alexanyan, Svitlana Tkachuk, Anatoliy Starodub, Taras Luchyshyn, Andriy Sibirny, Serhiy Souchelnytskyi, Yuriy Kit

## Abstract

Coronavirus disease 19 (COVID-19) is a global health crisis on a planetary scale. COVID-19 in many people has mild or moderate manifestation, although significant number of people, especially the elderly, suffer heavy from this illness, which often resulting in death. There are reports of similarities in immune response between COVID-19 and some autoimmune diseases. Earlier, we have demonstrated that fraction of TCA-soluble blood serum proteins containing a 48 kDA fragment of unconvential Myosin C1 have linked with development of multiple sclerosis and rheumatoid arthritis. Here we analyze use of these proteins in determining the severity of disease in COVID-19 patients. We found that blood serum of COVID-19 patients in acute disease manifestation contains, in contrast to healthy individuals, the TCA-soluble proteins with molecular masses 48 kDa and 76 kDA which were identified as a short form of unconventional myosin 1c and a modified form of human serum albumin.

## Background

Coronavirus disease 19 (COVID-19) is a global health crisis, where SARS-CoV-2 is the novel coronavirus responsible for this respiratory disease. COVID-19 has different manifestation. Majority of the infected people have only mild or moderate symptoms and recover without hospitalization. However, in a significant number of patients, especially the elderly, this disease causes pneumonia and thrombosis, which often leads to death. Establishing biomarkers for COVID-19 that indicate which patients are likely to develop severe disease could help clinicians to categorize patients earlier, improve prognosis and prioritize treatment [1-4]. Reported evidence suggests that some medications used to treat autoimmune rheumatologic diseases might have therapeutic effect in patients with severe COVID-19 infections, drawing attention to the relationship between COVID-19 and autoimmune diseases [5,6]. Recently it was demonstrated that autoantibodies are linked with SARS-CoV-2 infection could be the key to understanding some of the worst cases COVID-19 disease [7-10]. Summarized data suggested that autoimmune markers could be useful for estimation of disease severity of COVID-19 patients. Earlier we have found that high level of TCA-soluble proteins was observed in blood serum patients with multiple sclerosis and rheumatoid arthritis, and other autoimmune and lymphoproliferative diseases [11-13]. Here, we studied the presence of TCA-solubilized proteins in blood serum of COVID-19 patients with different ranges of severity. We observed that expression of these proteins may correlate with severity of COVID-19.

## Methods

This protocol in more detail is described below. The blood samples (2 ml) of were collected under the approval of the Bio-Ethics Review Board of the Danylo Halytsky Lviv National Medical University in accordance with the regulation of the Ministry of Health of Ukraine (number of the permit has to be added). A documented consent was obtained from all patients included in the study, and the form of the consent was approved by the Bio-Ethics Review Board of the Danylo Halytsky Lviv National Medical University. Blood serum was obtained after blood coagulation for 30 min at 23^0^C and followed centrifugation for 10 min at 5000 g. 200 μl of blood serum was diluted in 2-fold with phosphate buffer saline (PBS), and then 100% TCA was added to 10% of final concentration. After 30-min incubation on ice, the solution was centrifuged for 15 min at 10,000 g. 200 μl of the supernatant containing TCA-soluble compounds was transferred to a fresh Eppendorf tube (volume 0.15 ml) and cold acetone was added thereto to a final volume of 0.15 ml. Obtained mixture was incubation at −20 °C for 18 h and precipitate was pelleted by centrifugation for 30 min at 10,000 g. The pellet was dried at 37^0^C, diluted in 30 μl of distilled water and samples were stored at −20°C until use. The obtained samples were subjected to SDS-PAGE electrophoresis in 12% PAG [13], followed by staining of proteins with Coomassie brilliant blue G-250 in the gel. The individual electrophoretic polypeptide bands were excised from the gel and subjected to in-gel trypsin digestion, followed MALDI TOF/TOF mass spectrometry proteins identification according to [14].

## Results and Discussion

Earlier, various blood serum proteins of prognostic and diagnostic values were isolated and identified by us in blood serum using TCA-extraction in combination with mass-spectrometry [11, 12] (Fig. 1). Here, we used this approach to characterize the fraction of TCA-soluble proteins isolated from blood serum of the clinically healthy people and COVID-19 patients with severe disease. We compared the composition of the fractions of TSP, obtained from blood serum of 28 persons without any clinical disease manifestation, which were tested for anti SARS-CoV-2 antibody (IgG and IgM isotypes) in clinical diagnostic laboratory using Vitrotest® SARS-CoV-2 IgG and Vitrotest® SARS-CoV-2 IgM kits. (Fig 2 A). Among them, 4 persons were positive for anti SARS-CoV-2 IgGs (Fig.2 A lanes 1-4) and 2 persons were positive at SARS-CoV-2 IgM antibodies (Fig.2 A lanes 5-6) according to tests. Other 28 TCA-soluble blood serum protein samples belonged to COVID-19 patients, which were hospitalized due to a severe course of the disease (Fig. 2 B). Electrophoretic separation of TCA-soluble proteins isolated from the cohorts of clinical healthy people and patients with COVID-19 manifestation revealed essential difference of their proteins’ composition. It was found that the fractions of these proteins isolated from blood serum of healthy people mainly contain 66 kDa protein, while the same fraction isolated from blood serum of COVID-19 patients has strong expression of 48 kDa protein. Earlier, using the combination of high-performance liquid chromatography (HPLC), MALDI TOF/TOF mass spectrometry and Western blots we identified that a 66 kDa protein is a serum albumin, while 48 kDa protein have been identified as a N-terminal fragment of unconventional myosin 1c [11]. Needs to be noted that the fractions of TCA-soluble proteins, extracted from blood serum of COVID-19 patients also contained polypeptides with molecular mass 76 kDa (Fig. 2A). In order to identify these polypeptides, we used MALDI TOF/TOF mass spectrometry of 4 polypeptides isolated from spectra of electrophoretic bands of 4 patients, suffering with Covid-19 disease. The results obtained show that all 76 kDa polypeptides have, on average, 80% similarity to human serum albumin (Fig. 2B). We suggest that these polypeptides are modified (glycated or glycosylated) form of human blood serum albumin of original molecular mass 66 kDa. Origin of this modification needs to be additionally studied.

Comparative analysis of the composition of TCA-solubilized proteins revealed the presence of 48 kDa and 76 kDa proteins in blood serum of patients with acute COVID-19 manifestation versus clinical healthy people, where these polypeptides were not detected. The accumulated data convincingly demonstrate that 48 kDa polypeptide is a short form of unconvential myosin 1c earlier found by us in blood serum of patients with autoimmune and hematoncological disease [11,12]. It is known that Myo1C (molecular mass 121.7 kDa) is abundantly expressed in mammalian B lymphocytes [12] and the level of 48 kDa fragment in blood serum could be tightly associated with blood cells degradation [11,12,17]. Further investigation demonstrated that 48 kDa myosin 1C capable of binding in blood serum with IgG and IgM immunoglobulins [18], as well as with component 3 of the complement and the antithrombin-III [19]. Thus, we suggest that 48/Myo1C might be engaged in immunologic and coagulation cascades in the bloodstream of COVID-19 suffering patients.

Origination and functional activity of 76 kDa polypeptide revealed in fractions of TCA-solubilizes blood serum proteins of COVID-19 patients with acute disease manifestation needs further study. We suggest, that the presence of this blood serum protein might be linked to modification of monomeric (66 kDa) albumin in bloodstream of COVID-19 patients. Evidence suggests that these modifications could result in oxidative stress observed in some healthy people [20, 21] as well as in patients with diabetes [22].

## Conclusion

Summarized data demonstrate that blood serum of COVID-19 patients in acute disease manifestation contains, in contrast to healthy individuals, the TCA-soluble proteins with molecular masses 76 and 48 kDa which were identified as a short form of unconventional myosin 1c and a modified form of human serum albumin. We suggest that obtained data provide a basis for further in-depth studies of the detected proteins in development of COVID-19.

## Data Availability

All data is available

https://www.researchgate.net

## Abbreviation

TCA: 2,2,2-trichloroacetic acid

---

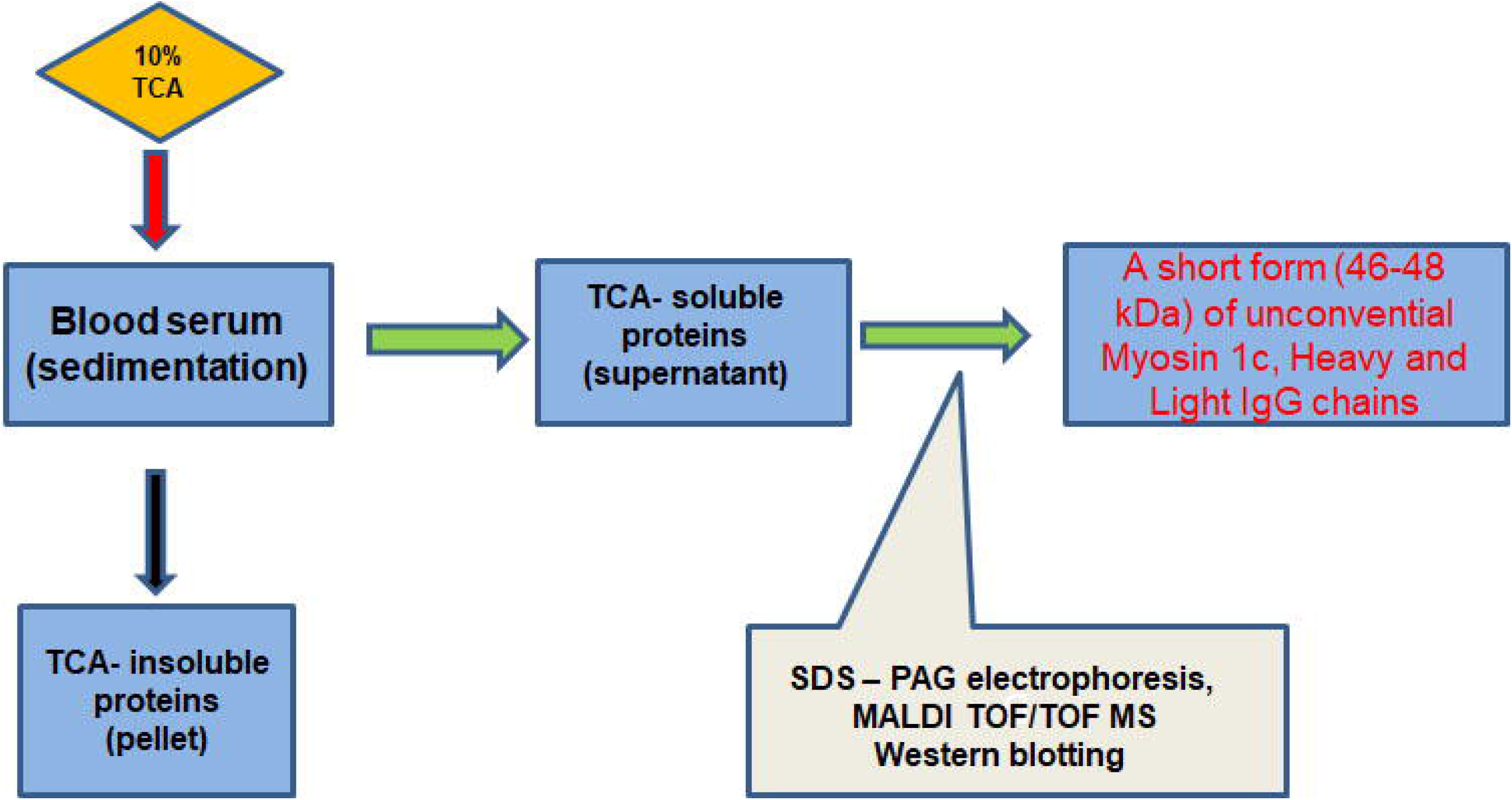

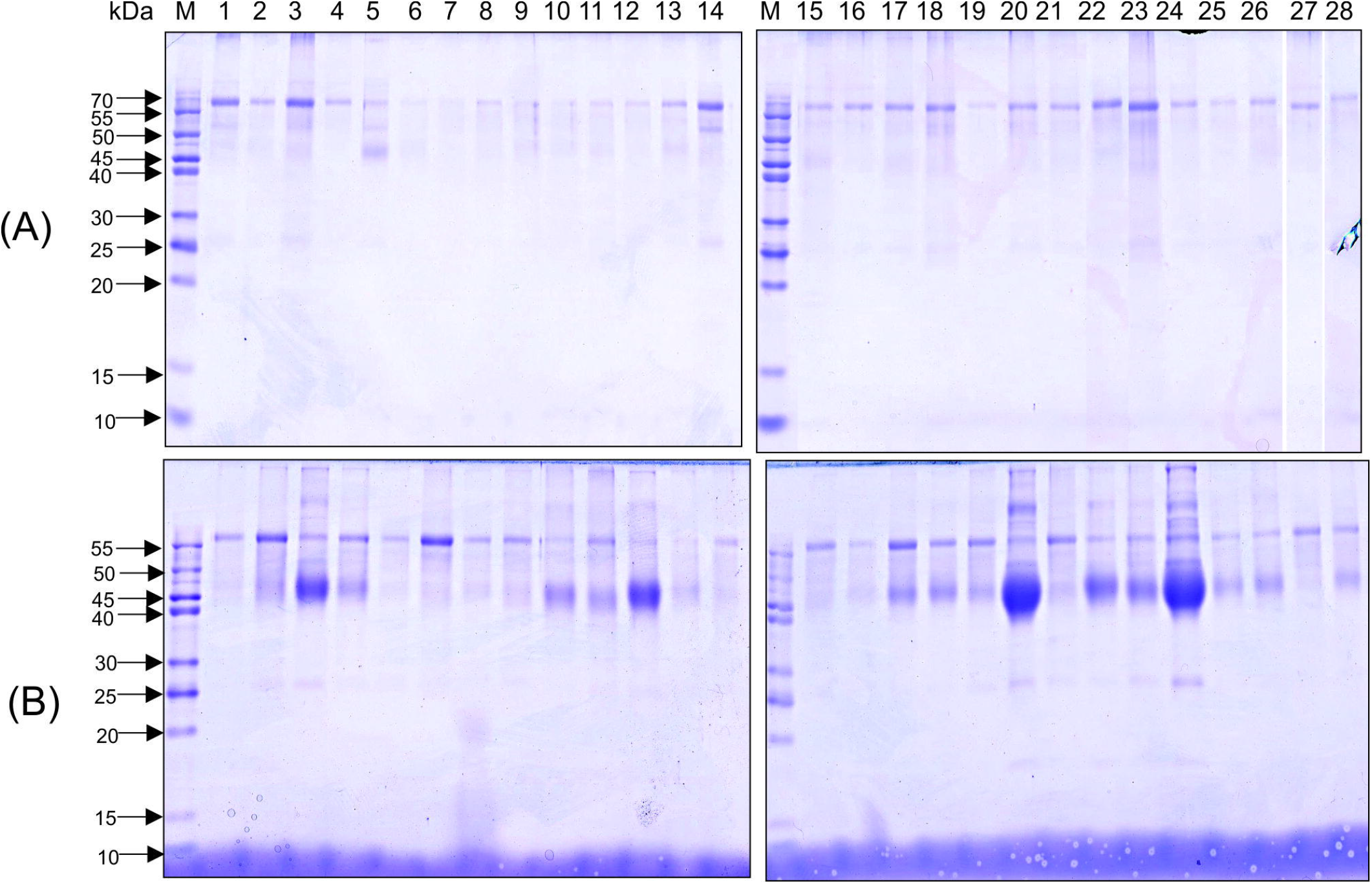

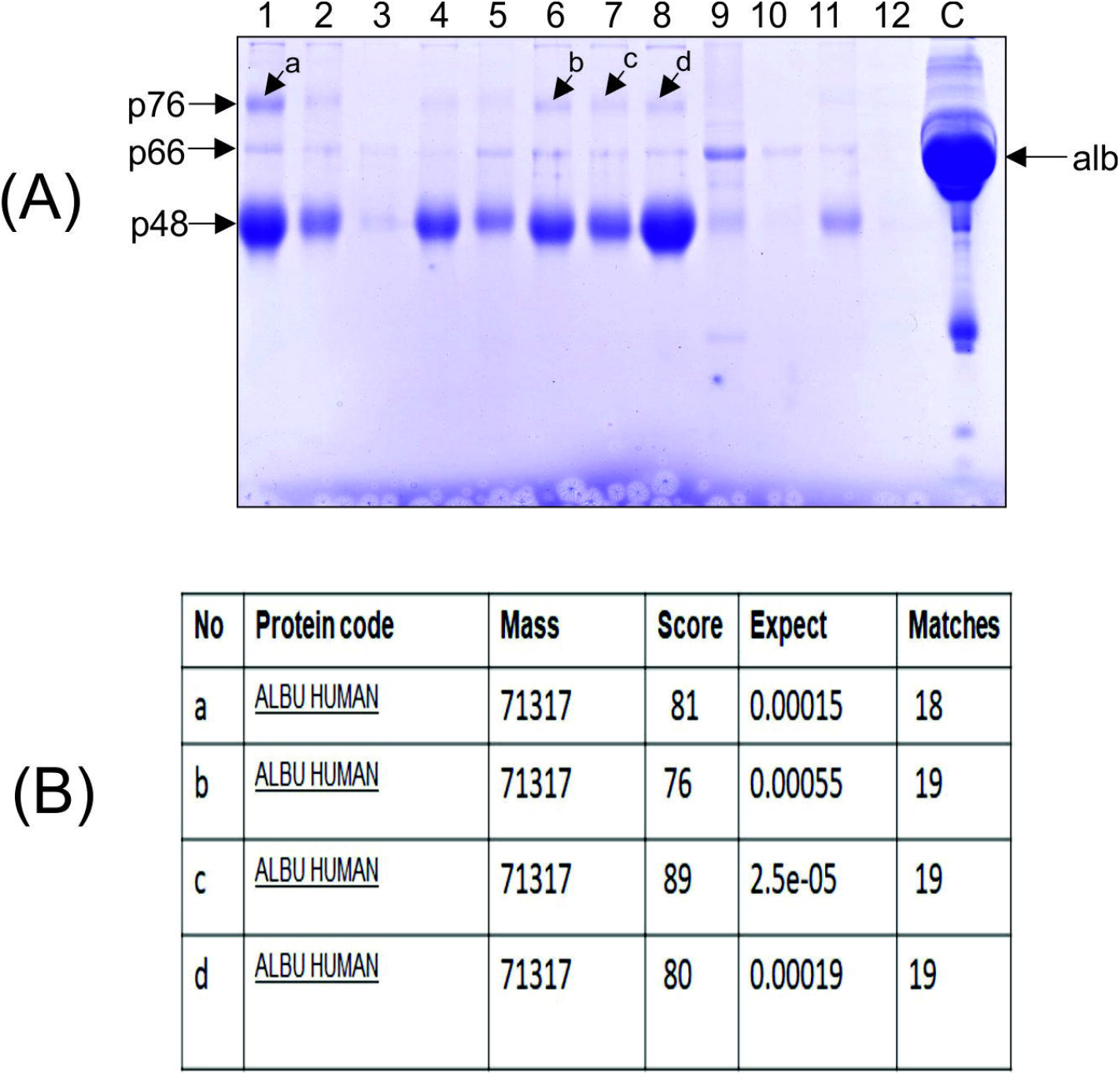

